# Degradation of bleach produced for disinfection in Kenyan healthcare facilities using novel technology

**DOI:** 10.64898/2026.08.03.26359620

**Authors:** Jared Oremo, Sunkyung Kim, Alex Mwaki, Robert Quick

**Affiliations:** Safe Water and AIDS Project, P.O.Box 3323, 40100 Kisumu, Kenya; Independent statistical consultant, Atlanta, GA, USA

**Keywords:** Bleach, bleach degradation, disinfection, healthcare facilities, hypochlorite generator

## Abstract

Disinfection with bleach is recommended for surface cleaning in healthcare facilities, but bleach degrades over time, reducing disinfection effectiveness. We evaluated whether local bleach produced by hospitals using novel technology had adequate shelf life to justify use. Two hospitals (A and B) each produced and stored 0.5% bleach in two 20-liter plastic containers; bleach in one container was alkalinized for increased stability to pH 12; bleach in the other container was not stabilized and, for comparison, a third container of commercial bleach was tested. We tested three bleach samples from each of the containers produced by hospitals A and B, and commercial bleach for free available chlorine (FAC) using N,N-diethyl-p-phenylenediamine at irregular intervals up to 216 days. We compared the expected percent decrease in FAC per 7 days by bleach type using log-normal regression. Stabilized bleach decreased by 1% and 2% in hospitals A and B, respectively, every 7 days while non-stabilized bleach decreased by 21% and 12%, respectively (all p<0.001); commercial bleach decreased by <1% per 7 days. Bleach production proved feasible in hospitals, and both stabilized and commercial bleach maintained adequate concentration for disinfection; non-stabilized bleach maintains a useful concentration for not longer than three weeks after production.

## Introduction

Inadequate infection prevention and control practices, including lack of access to basic water, sanitation, and hygiene (WASH) infrastructure, in healthcare facilities (HCFs) are serious problems in developing countries, posing risks to the health of patients and healthcare providers (Guo, *et al*., 2017, Cronk, *et al*., 2018, Haque, *et al*., 2020) . WASH improvements in developing country HCFs can help prevent healthcare-acquired infections, and surface disinfection is an important WASH component (Haque, *et al*., 2020). Although disinfection with sodium hypochlorite is one recommended approach (Haque, *et al*., 2020), bleach supplies are often inadequate in developing country settings because of budget shortfalls (Yadav 2015), inadequate supply chains (Yadav 2015), poor-quality commercial bleach (Lantagne 2009), and degradation of bleach over time, which can reduce disinfection effectiveness (Ogutu, *et al*., 2001, Helmenstine 2021).

To address sodium hypochlorite supply issues for HCFs, a variety of hypochlorite generators have been developed to permit local bleach production and distribution. For this evaluation, we used the STREAM disinfectant generator manufactured by Aqua Research (Albuquerque, NM, USA) to produce 0.5% sodium hypochlorite solution electrolytically from a 15-grams/liter brine solution (Fig 1). The STREAM device is designed to be portable; can operate on 110 or 220 volt, 50 to 60 Hz power, a 12-volt automobile battery, or solar panels; is housed in a waterproof hard plastic case; is manufactured with waterproof components; has a control mechanism to ensure the correct output bleach concentration of 5000 mg/l (0.5%) even with improperly made brine solution or if there are intermittent power outages during chlorine production; and can be effectively cleaned using a vinegar solution. One recent enhancement of the STREAM device is an automatic data collection and GPS location system that can be Bluetooth-connected to the internet to permit remote monitoring of operation and maintenance data.

**Figure 1.**
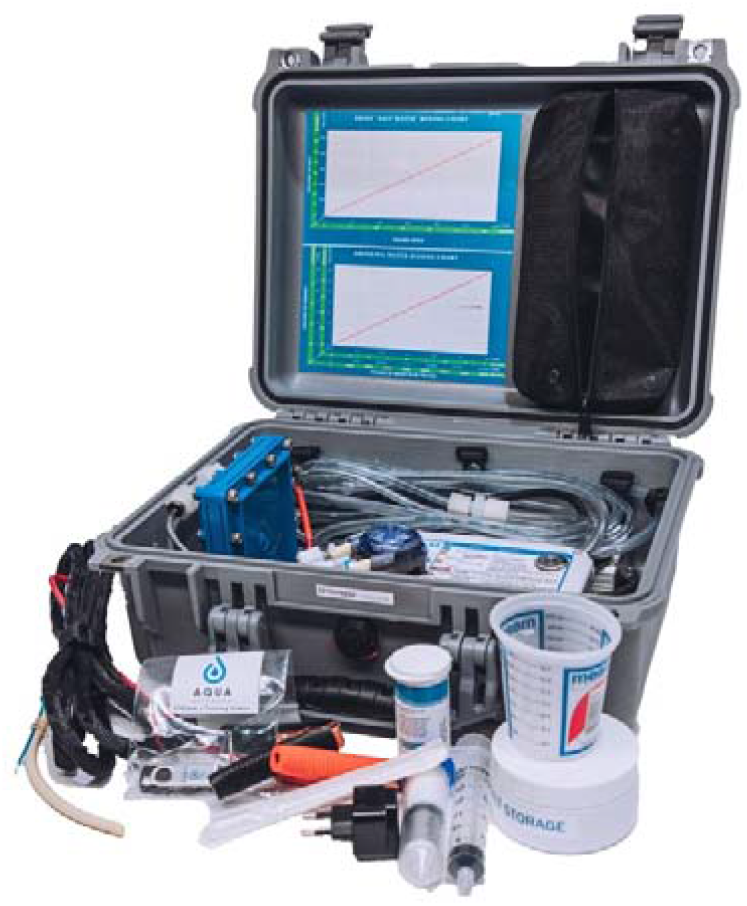
Photograph of STREAM device

The shelf life of bleach is relatively short (Ogutu, *et al*., 2001, Helmenstine 2021), which can limit its usefulness in health care settings. However, if bleach is alkalinized to a pH of 11 or 12, shelf life is increased substantially (Centers for Disease Control and Prevention 2022). To assess the shelf life of STREAM-generated bleach solution – and, therefore, its utility in HCFs - Aqua Research partnered with the Safe Water and AIDS Project (SWAP – a Kenyan NGO), and the Kisumu County Government’s Department of Health and Sanitation to deploy STREAM devices in two hospitals in western Kenya and compare the bleach degradation rate in alkalinized and non-alkalinized samples over a 6-month period.

## Materials and Methods

A STREAM device was installed in hospital A on 25 February 2022 and in hospital B on 15 March 2022. Hospital personnel were trained in the production of chlorine, and operation and maintenance of the devices. To evaluate the shelf life of sodium hypochlorite solution (hereafter referred to as bleach) produced by the STREAM devices, we requested hospitals A and B to generate bleach and store it in two 20-liter black containers, one designated for stabilized bleach and the other for non-stabilized bleach. SWAP personnel traveled to each hospital, transported the bleach containers to the SWAP water laboratory, and stored them at ambient temperature. For comparison purposes, three one-liter containers of commercial sodium hypochlorite was purchased for each hospital and stored under the same conditions in the SWAP laboratory.

To prepare the bleach samples for testing, we filled three one-liter bottles from each of the two 20-liter containers of STREAM bleach (one stabilized, one non-stabilized) obtained from hospitals A and B as well as obtained three one-liter containers of commercial bleach per hospital, for a total of 18 one-liter bottles. To prepare stabilized bleach samples, we set aside three bottles from each of the 20-liter containers of STREAM bleach from hospitals A and B designated for stabilized bleach, for a total of six bottles. To these six bottles, we added sodium hydroxide pellets in increments of one gram, dissolving the pellets in the bleach, measuring the pH with a Starter 3100 Bench pH Meter (Ohaus Corp., Parsippany, NJ), and repeating the process until we obtained the target pH of 11.5 to 12. The six bottles of STREAM bleach designated as non-stabilized and the six one-liter bottles of commercial bleach were stored untreated. We tested the bleach in each of the 18 bottles for temperature using a Thermo Scientific Eutech PC Tester Multiparameter Pocket Tester (Fisher Scientific Co, Hampton, NH), for pH as noted above, and for free available chlorine (FAC) on the day of arrival at the SWAP laboratory (day 2 following production for Hospital A and day 0 for Hospital B) using a Hach Pocket Colorimeter (Hach Co, Loveland, CO) (see Tables 1 and 2). To measure FAC, we pipetted three one-ml samples from each of 18 bottles and added each of the samples to beakers filled with 999ml of chlorine-demand-free diluent and mixed them well. We then pipetted 10 ml of solution from each of 54 beakers into test tubes, dissolved N,N-diethyl-p-phenylenediamine (DPD) reagent in each of the 54 test tubes, and determined FAC concentration in parts per million (ppm).

**Table 1.** Multiplicative decrease in FAC per 7 days, by bleach type and hospital, Kenya, 2022.

|  | Hospital A |  |  | Hospital B |  |  |
| --- | --- | --- | --- | --- | --- | --- |
|  | Estimate | 95% CI | P-value | Estimate | 95% CI | P-value |
| Non-stabilized | 0.848 | 0.822, 0.874 | $<0.001$ | 0.913 | 0.912, 0.914 | $<0.001$ |
| Stabilized | 0.992 | 0.992, 0.992 | $<0.001$ | 0.987 | 0.986, 0.987 | $<0.001$ |
| Commercial | 0.998 | 0.998, 0.998 | $<0.001$ | 0.997 | 0.997, 0.998 | $<0.001$ |

To examine the rate of bleach degradation, we tested the samples, using identical procedures to those described above, at approximate intervals of one to three times per week for up to 2 months, then approximately once a month thereafter. The irregular intervals resulted from attempts to increase efficiency by harmonizing the timing of testing of samples from the two hospitals (which had different start dates), and because of uneven availability of testing supplies and personnel in the laboratory, and a need to replace mother boards at different times in the two STREAM devices. Testing continued through day 216 in hospital A and day 196 in hospital B.

FAC results of the three specimens from each bottle were averaged before analysis. We assessed and compared the expected percent degradation of bleach in ppm per 7 days between non-stabilized and stabilized bleach by applying multivariable log-normal regression, treating log-transformed FAC as a dependent variable and bleach type and testing day as independent variables with an interaction effect of bleach type and testing day. The potential correlation between the FAC measures from the same bottle over multiple testing days was considered using generalized estimating equation approach. For commercial product, simple log-normal regression was applied. All analyses were run separately by hospital.

## Results

During the period of this evaluation, hospital A produced approximately 3520 liters of bleach and hospital B produced approximately 4550 liters. STREAM devices at each hospital were down for approximately one month during the evaluation period to obtain and replace the motherboards.

In Hospital A, no data were obtained at time 0 because the bleach containers were first received at the SWAP laboratory two days after production. The temperature of bleach samples from hospital A tested during this evaluation ranged from 24 to 30°C. The mean FAC of non-stabilized bleach produced at hospital A ranged from 5200 ppm on day two to 167 ppm by day 216 (decrease of 96.8%) (Figure 2); FAC remained above 4000 ppm through day 22. Solution pH ranged from 7.2 to 9.0. The mean FAC of stabilized bleach produced at Hospital A was 5278 ppm on day 2 and gradually decreased to 4344 ppm by day 216 (decrease of 18%); FAC remained above 5000 ppm through day 29. During the evaluation, pH ranged from 11.0 to 12.8. The mean FAC of commercial bleach from Hospital A was 33,556 ppm on day 2 and gradually decreased to 31,889 ppm by day 216 (decrease of 5%). During the evaluation, pH ranged from 11.9 to 13.3.

**Figure 2.**
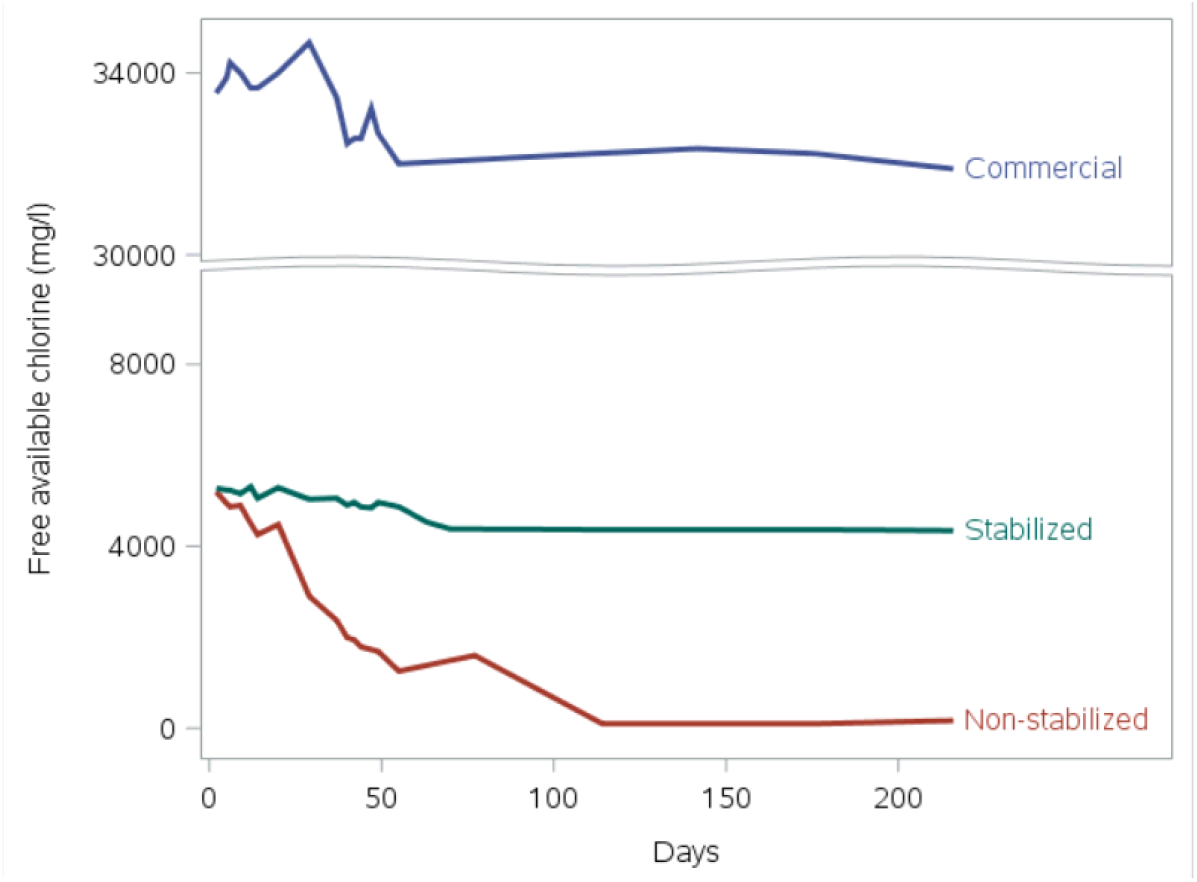
Mean FAC by days following production by bleach type, Hospital A, Kenya, 2022

The temperature of bleach solution samples from hospital B tested during this evaluation ranged from 24 to 27°C. The mean FAC of non-stabilized bleach produced at hospital B ranged from 6144 ppm on the day of production to 422 ppm by day 196 (decrease of 93.1%); FAC remained above 4000 ppm through day 44 (Figure 3). Solution pH ranged from 7.4 to 9.0. The mean FAC of stabilized bleach produced at Hospital B was 6267 ppm on the day of production, and gradually decreased to 4411 ppm by day 196 (decrease of 29.6%); FAC remained above 5000 ppm through day 29. Solution pH ranged from 11.0 to 12.8. The mean FAC of commercial bleach from hospital B was 33,889 ppm on day 2 and gradually decreased to 31,889 ppm by day 196 (decrease of 5.9%). During the evaluation, pH ranged from 11.5 to 12.5.

**Figure 3.**
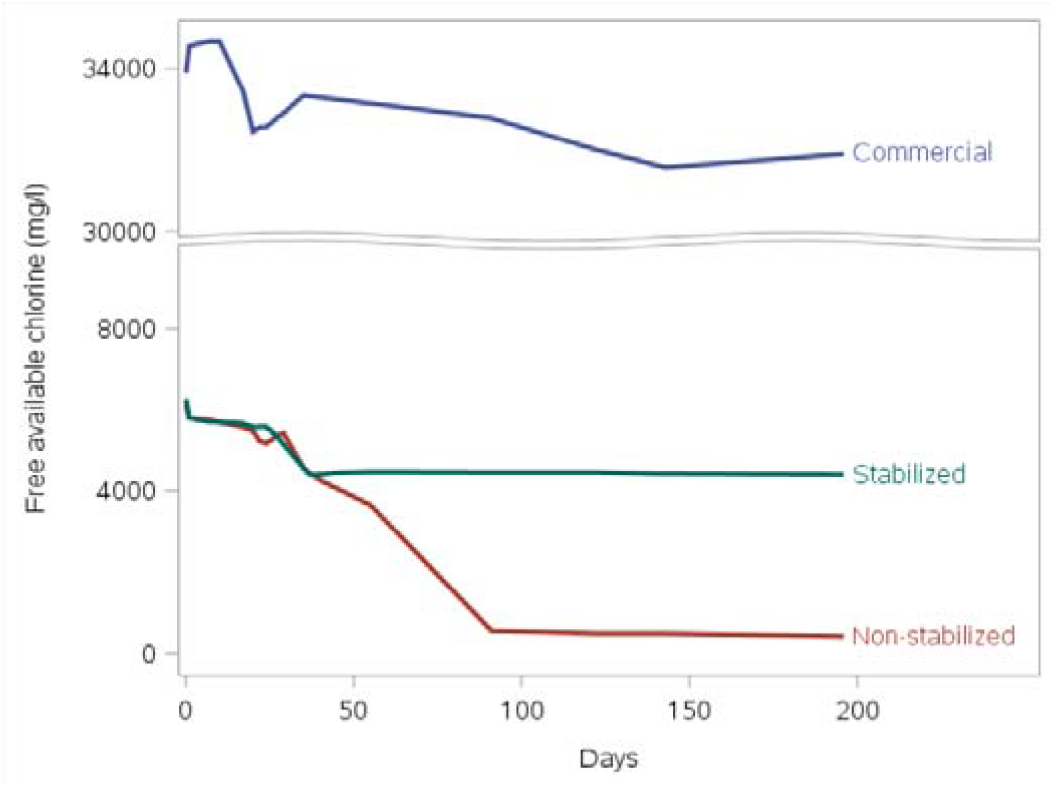
Mean FAC by days since production by bleach type. Hospital, Kenya, 2022

In hospital A, every 7 days, FAC decreased by 21% (multiplicative effect or ME in FAC: 0.789) for non-stabilized bleach and by 1% (ME=0.986) for stabilized bleach (all p<0.001) (Table 1). In hospital B, FAC decreased by 12% (ME=0.878) for non-stabilized and by 3% (ME=0.974) for stabilized bleach per 7 days (all p<0.001) (Table 1). In both hospitals, the commercial product decreased by <1% (p<0.001) per 7 days.

In hospital A, the multiplicative decrease in FAC per 7 days for non-stabilized bleach was significantly greater than the multiplicative decrease in FAC per 7 days for stabilized bleach (p=0.027) and commercial bleach (p=0.015). In hospital B, the multiplicative decrease in FAC per 7 days for non-stabilized bleach was significantly greater than the multiplicative decrease in FAC per 7 days for stabilized bleach (p=0.015) and commercial bleach (p=0.024).

## Discussion

Results of this evaluation demonstrated that bleach produced by the STREAM device has a shelf life of at least six months if alkalinized to a pH of approximately 12. The FAC concentration of stabilized bleach decreased by 17% to 29% during the evaluation period, remaining well above 4000 ppm, and was still above 5000 ppm in both hospitals during the first month after production. At this concentration, the STREAM bleach would be adequate for most hospital disinfection needs (Haque, *et al*., 2020). For surface disinfection or instrument sanitization, a concentration of 1000 ppm is recommended (World Health Organization 2020), requiring a dilution of 5:1 or 4:1 (depending on concentration at the time of dilution). Although a bleach concentration of 5000 ppm is recommended for cleaning blood or body fluid spills, the recommendation is to first remove organic material by cleaning the spill with detergent and water, then disinfecting with bleach (World Health Organization 2020). The concentration of pH-stabilized STREAM bleach measured in this evaluation should be adequate for this purpose, particularly during the first month after production. Because commercial bleach would need to be diluted by a factor of 33:1, an additional advantage of STREAM bleach compared to commercial bleach is the relative greater simplicity of a 5:1 or 4:1 dilution. If fresh STREAM bleach was produced and completely used up within one to four weeks, then stabilization of the bleach would not be necessary.

An additional potential benefit of local production of bleach is its application to drinking water (Clasen, *et al*., 2007). To obtain a concentration of approximately 2mg/l, which can provide palatable, potable water would require a dilution of 2000:1, which would require approximately 8mg (approximately 1.5 teaspoons) of bleach in 20 liters of water.

Although compared to commercial bleach, the concentration of alkalinized STREAM bleach appeared somewhat less stable over time, its effectiveness would likely be similar to the commercial product. Programs to provide STREAM bleach to HCFs should encourage liberal use of the disinfectant to ensure effectiveness and use up the bleach while its concentration is optimal for disinfection. Despite the relatively lower stability of STREAM bleach compared to the commercial product, the convenience and reliability of local production offer certain advantages, including mitigation of supply chain issues (Yadav 2015), lower commodity cost because the only inputs are salt, water, and electricity; and reliable concentration (Lantagne 2009, Helmenstine 2021). During this study, both STREAM devices produced over 3500 liters of bleach, demonstrating their potential for producing sufficient quantities for the hospitals with leftover volume that would be available for distribution to surrounding HFCs. Currently, 207 STREAM devices have been installed in healthcare facilities in 10 African countries and Haiti (Personal communication, Chris Dunston, Director of Partnerships, Marketing, and Sales, Aqua Research, Inc), which offers abundant opportunities for further study of this potential benefit as well as cost benefit analyses.

This study had several important limitations. First, because the project took place in a convenience sample of two hospitals in one county in western Kenya, the results are not generalizable. Second, bleach FAC was tested at irregular intervals because of logistical issues. However, because the main evaluation question concerned bleach degradation over time, slight differences in testing intervals present no problem in assessing bleach shelf life, with the relatively high number of data points smoothing out the degradation curve. Third, at the time of preparation of this manuscript, the evaluation had been underway for over six months, limiting the assessment of bleach degradation to a relatively short period. However, program objectives include monthly distribution of fresh bleach and encouragement to use the product liberally. Bleach shelf life documented in this evaluation is sufficient to meet program objectives. Fourth, the up-front cost of the STREAM device (approximately $4,000) presents a financial barrier to program initiation that will likely require funding from non-governmental sources to succeed. Finally, quality control of solution will require reagents and laboratory supplies that will need to be periodically replenished, and operator training, all of which have costs.

## Conclusions

In conclusion, local production of bleach in Kenyan hospitals using a portable hypochlorite generator was feasible. Although non-stabilized bleach maintained its concentration for three to four weeks following production, stabilization of the solution to a pH of 11 to 12 will extend the shelf life to six months or more and is recommended.

## Data Availability

All data produced in the present study are available upon reasonable request to the authors.

## Supplementary Materials

Table S1: Temperature, pH, and average free available chlorine (FAC) for stabilized and non-stabilized STREAM bleach, and commercial bleach, by number of days following production, Hospital A, Western Kenya, 2022; Table S2: Temperature, pH, and average free available chlorine (FAC) for stabilized and non-stabilized STREAM bleach, and commercial bleach, by number of days following production, Hospital B, Western Kenya, 2022.

## Data availability statement

All data from this evaluation are contained in Supplementary tables 1 and 2.

## Acknowledgments

We are grateful for the support of SWAP staff and the SWAP technical advisor, Alie Eleveld.

