## Supplementary tables 1 and 2 for "Degradation of bleach produced for disinfection in Kenyan healthcare facilities using novel technology"

Supplementary table 1. Temperature, pH, and average free available chlorine (FAC) for stabilized and non-stabilized STREAM bleach, and commercial bleach, by number of days following production, Hospital A, Western Kenya, 2022

| Days from production | Non-stabilized | | | | Stabilized | | | | Commercial | | | |
| --- | --- | --- | --- | --- | --- | --- | --- | --- | --- | --- | --- | --- |
|  | Temp | Ph | FAC | Range | Temp | Ph | FAC | Range | Temp | Ph | FAC | Range |
| 0† | NA | | | | | | | | | | | |
| 2 | . | 9.2 | 5200 | 5100-5300 | . | 12.0 | 5278 | 5100-5400 | . | 12.1 | 33556 | 32000-35000 |
| 5 | 25.1 | 8.8 | 4944 | 4800-5100 | 25.0 | 11.9 | 5233 | 5100-5400 | . | 12.1 | 33889 | 33000-35000 |
| 6 | 25.0 | 8.7 | 4867 | 4800-5000 | 25.1 | 11.7 | 5233 | 5000-5400 | 25.0 | 12.0 | 34222 | 34000-35000 |
| 9 | 25.0 | 9.5 | 4900 | 4800-5000 | 29.0 | 12.3 | 5156 | 5000-5400 | 29.0 | 12.5 | 34000 | 33000-36000 |
| 12 | 25.0 | 9.0 | 4500 | 4400-4600 | 26.0 | 12.6 | 5311 | 5200-5400 | 25.2 | 12.4 | 33667 | 33000-34000 |
| 14 | 26.1 | 8.7 | 4256 | 4100-4400 | 25.0 | 12.6 | 5056 | 4900-5200 | 25.0 | 12.3 | 33667 | 33000-34000 |
| 20 | 29.8 | 9.3 | 4478 | 4300-4600 | 29.9 | 12.8 | 5289 | 5200-5400 | 30.2 | 13.3 | 34000 | 33000-35000 |
| 29 | 24.0 | 8.0 | 2900 | 2300-3200 | 25.0 | 12.2 | 5033 | 5000-5100 | 25.0 | 12.8 | 34667 | 34000-35000 |
| 37 | 25.0 | 7.9 | 2367 | 1700-2800 | 24.1 | 12.2 | 5056 | 4900-5200 | 25.1 | 12.9 | 33444 | 33000-34000 |
| 40 | 24.0 | 7.9 | 1989 | 1400-2400 | 25.0 | 12.3 | 4900 | 4800-5000 | 25.1 | 12.8 | 32444 | 32000-33000 |
| 42 | 24.0 | 7.8 | 1944 | 1400-2300 | 24.2 | 12.2 | 4967 | 4900-5000 | 25.0 | 12.6 | 32556 | 32000-33000 |
| 44 | 24.0 | 7.8 | 1789 | 1300-2200 | 25.1 | 12.2 | 4867 | 4700-5000 | 24.4 | 12.8 | 32556 | 32000-33000 |
| 49 | 23.9 | 7.2 | 1689 | 1500-1900 | 25.0 | 11.5 | 4967 | 4800-5100 | 24.0 | 12.5 | 32667 | 32000-33000 |
| 55 | 25.0 | 7.2 | 1256 | 900-1600 | 24.0 | 11.5 | 4867 | 4800-4900 | 24.0 | 12.4 | 32000 | 31000-33000 |
| 142 | 24.1 | 7.4 | 100 | 100-100 | 23.3 | 11.0 | 4367 | 4200-4500 | 23.7 | 12.4 | 32333 | 31000-33000 |
| 176 | 23.1 | 7.4 | 100 | 0-200 | 24.2 | 11.0 | 4367 | 4200-4500 | 23.6 | 12.3 | 32222 | 31000-33000 |
| 216 | 24.0 | 7.4 | 167 | 100-300 | 24.1 | 11.0 | 4344 | 4200-4400 | 24 | 12 | 31889 | 31000-34000 |

†Bleach was not tested until day 2 following production

Supplementary table 2. Temperature, pH, and average free available chlorine (FAC) for stabilized and non-stabilized STREAM bleach, and commercial bleach, by number of days following production, Hospital B, Western Kenya, 2022

| Days from production | Non-stabilized | | | | Stabilized | | | | Commercial | | | |
| --- | --- | --- | --- | --- | --- | --- | --- | --- | --- | --- | --- | --- |
|  | Temp | Ph | FAC | Range | Temp | Ph | FAC | Range | Temp | Ph | FAC | Range |
| 0 | 26.9 | 9.0 | 6144 | 5900-6300 | 25.0 | 12.8 | 6267 | 6100-6400 | 25.0 | 12.5 | 33889 | 33000-35000 |
| 1 | 25.0 | 8.7 | 5800 | 5700-5900 | 25.1 | 12.6 | 5800 | 5800-5800 | 25.0 | 12.5 | 34556 | 34000-36000 |
| 7 | 26.0 | 8.6 | 5767 | 5700-5800 | 25.1 | 12.6 | 5700 | 5700-5700 | 26.0 | 12.4 | 34667 | 34000-35000 |
| 10 | 25.2 | 8.4 | 5689 | 5600-5700 | 25.1 | 12.6 | 5700 | 5700-5700 | 26.8 | 12.3 | 34667 | 34000-35000 |
| 17 | 26.2 | 8.0 | 5567 | 5400-5700 | 26.0 | 12.4 | 5667 | 5600-5700 | 26.2 | 12.0 | 33444 | 33000-34000 |
| 20 | 26.4 | 7.8 | 5478 | 5400-5600 | 26.2 | 12.2 | 5567 | 5500-5600 | 26.0 | 12.0 | 32444 | 32000-33000 |
| 22 | 26.0 | 7.8 | 5222 | 5000-5400 | 26.0 | 12.0 | 5578 | 5500-5700 | 25.8 | 12.0 | 32556 | 32000-33000 |
| 24 | 26.8 | 7.8 | 5178 | 5000-5300 | 26.2 | 12.0 | 5578 | 5500-5700 | 26.0 | 12.0 | 32556 | 32000-33000 |
| 27 | 24.0 | 7.8 | 5356 | 5300-5500 | 25.0 | 11.9 | 5356 | 5300-5500 | 24.0 | 12.0 | 32778 | 30000-34000 |
| 29 | 25.0 | 7.8 | 5433 | 5300-5500 | 26.0 | 12.0 | 5156 | 5100-5300 | 24.0 | 12.2 | 32889 | 30000-34000 |
| 35 | 25.0 | 7.7 | 4578 | 4400-4900 | 25.0 | 11.8 | 4578 | 4400-4900 | 25.0 | 12.0 | 33333 | 33000-34000 |
| 91 | 25.5 | 7.8 | 567 | 400-900 | 26.0 | 11.5 | 4467 | 4400-4600 | 25.0 | 12.2 | 32778 | 30000-34000 |
| 122 | 25.1 | 7.8 | 500 | 400-800 | 24.0 | 11.5 | 4467 | 4400-4600 | 25.2 | 12.2 | 32000 | 32000-32000 |
| 143 | 25.4 | 7.6 | 500 | 400-600 | 26.0 | 11.4 | 4433 | 4300-4600 | 25.0 | 11.8 | 31556 | 31000-32000 |
| 196 | 24 | 7.5 | 422 | 300-500 | 24 | 11.3 | 4411 | 4300-4500 | 24.0 | 12.0 | 31889 | 31000-32333 |
